# Decision for hysterectomy as a modifiable cardiovascular risk factor: A systematic review and meta-analysis

**DOI:** 10.64898/2026.11.11.26367416

**Authors:** DMCS Jayasundara, TDKM Jayasingha, IA Jayawardane, S De Silva Weliange, IU Ratnasekera

**Author notes:** Corresponding author: (DMCSJ).

## Abstract

**Objective:** To describe the role of hysterectomy in modifying the major atherosclerotic cardiovascular disease risk factors.

**Data sources:** PubMed, Cochrane Library, Embase, and ScienceDirect were searched from database inception to March 10, 2026.

**Study eligibility criteria:** Full-text randomized controlled trials, case-control studies, cross-sectional studies, cohort studies, case series, and experimental studies reporting new-onset cardiovascular events, diabetes mellitus, hypertension, dyslipidemia, or other metabolic effects following hysterectomy were eligible. Reviews and book chapters were excluded.

**Study appraisal and synthesis methods:** Study selection and data extraction were performed independently by multiple reviewers with conflicts resolved by consensus. Cohort study quality was assessed with the Newcastle-Ottawa Scale; a modified version assessed cross-sectional studies. Meta-analyses were performed using a generic inverse-variance random-effects model for each outcome, stratified by study design (cross-sectional versus cohort) and effect measure (adjusted odds ratio, adjusted hazard ratio, or relative risk). Narrative synthesis was performed for outcomes not meeting criteria for meta-analysis.

**Results:** Twenty studies (11 cohort, 8 cross-sectional, 1 mixed design) were included after screening 18,337 records from four databases. Hysterectomy was associated with significantly increased risk of hypertension (cross-sectional odds ratio 1.32, 95% confidence interval 1.18– 1.48; cohort hazard ratio 1.19, 95% confidence interval 1.09–1.30), diabetes (cross-sectional odds ratio 1.21, 95% confidence interval 1.11–1.32; cohort hazard ratio 1.20, 95% confidence interval 1.13–1.29), hyperlipidemia (hazard ratio 1.21, 95% confidence interval 1.13–1.29), obesity (odds ratio 1.24, 95% confidence interval 1.03–1.50), and metabolic syndrome (odds ratio 1.24, 95% confidence interval 1.08–1.44).

**Conclusions:** Hysterectomy is associated with increased risk of hypertension, diabetes, hyperlipidemia, obesity, and metabolic syndrome, key modifiable atherosclerotic cardiovascular disease risk factors. Active cardiovascular risk assessment and follow-up may be warranted for women who undergo hysterectomy, particularly at a young age.

## Introduction

Hysterectomy is the surgical removal of the uterus [1]. It is one of the most common gynecological procedures, with estimates of 0.6 million women undergoing hysterectomy annually in the United States [2]. Around 90% of surgeries are conducted to manage benign gynecological conditions like abnormal uterine bleeding, endometriosis, and uterine fibroids, where approximately 44% undergo concomitant bilateral salpingo-oophorectomy as part of the treatment process or to mitigate the risk of ovarian malignancy [3].

Although the treatment approaches for benign gynecological conditions have significantly evolved, hysterectomies are still increasingly performed owing to their low perioperative morbidity and rapid therapeutic effects [4]. Emerging evidence from recent systematic reviews and meta-analyses suggests a positive epidemiological association between hysterectomy (even with ovarian conservation) and increased long-term cardiovascular disease risk [4–6]. However, the data don’t emphasize specific pathophysiological mechanisms, making it difficult to distinguish chance from causation [4,5].

Cardiovascular risk factors are multifactorial, and some are modifiable [7–10]. Puri et al. classify cardiovascular risk factors as major atherosclerotic cardiovascular disease (ASCVD) risk factors, high-risk features, and risk modifiers [8]. Major ASCVD risk factors include age ≥ 45 in males and ≥ 55 in females, current cigarette smoking or tobacco use, and low high-density lipoprotein cholesterol (HDL) [8]. Family history of premature ASCVD, chronic kidney disease stage 3B or 4, apolipoprotein B > 130 mg/dl, extreme elevation of a single risk factor, lipoprotein (a) ≥ 50 mg/dl, metabolic syndrome, non-alcoholic fatty liver disease with grade two or three fibrosis, and a coronary artery calcium score < 75th percentile are among high-risk features [7,8]. Risk modifiers include lipoprotein a 20-40 mg/dl, impaired fasting glucose (100-125 mg/dl), increased waist circumference (> 90 cm in men and > 80 cm in women), high sensitivity C-reactive protein > 2mg/L, plasma triglyceride > 150 mg/dl (fasting), rheumatoid arthritis, psoriasis, spondyloarthritis, premature menopause, pre-eclampsia, gestational diabetes, polycystic ovarian syndrome, high polygenic risk score, air pollution and human immunodeficiency virus [8,10].

### Objective

To comprehensively evaluate the association between hysterectomy and cardiovascular risk, we need to establish the correlation between hysterectomy and the well-established cardiovascular risk factors. As it was not feasible to analyze every risk factor within the categories listed above, we selected five main risk factors in accordance with the European Society of Cardiology (ESC) guidelines [7,9,10]. If these five show a stronger association, we can then analyze additional factors. ESC guidelines describe the main causal and modifiable ASCVD risk factors as cholesterol (apo-B-containing lipoproteins), hypertension, cigarette smoking, diabetes mellitus, and adiposity [9]. To the best of the authors’ knowledge, this is the first systematic review and meta-analysis to synthesize evidence on the role of hysterectomy in modifying the main modifiable ASCVD risk factors. However, a long process of data exploration, synthesis, and analysis precedes classifying hysterectomy as a ‘high-risk feature’ or ‘risk modifier’. This review will ultimately shift how hysterectomy is viewed, from a gynecological procedure to a broader medical intervention.

## Materials and methods

### Information sources

We retrieved the relevant studies from PubMed, Cochrane Library, Embase, and ScienceDirect, published up to March 10, 2026.

### Search strategy

The search strings included “Hysterectomy”, “Cardiovascular disease”, “Coronary artery disease”, “Heart disease”, “Ischemic heart disease”, “Cardiovascular risk”, “Cardiac morbidity”, “Coronary heart disease”, “Diabetes”, “Hypertension”, “Dyslipidemia”, “Hyperlipidemia”, “Metabolic syndrome”, “Metabolic morbidity” which were combined using Boolean expressions “AND,” “OR” to form precise search queries. A detailed line-by-line search strategy is given in S1 File.

### Study selection

The study selection process was conducted in a blinded, two-step approach using the semi-automated tool Rayyan [11], involving two independent reviewers (TDKMJ) and (DMCSJ). In the first round, title and abstract screening removed duplicates, with conflicts resolved by (SDS); similarly, the second round used a similar blinded approach for full-text screening, with conflicts resolved by (AJ). The study selection process followed the PRISMA 2020 flow diagram for updated systematic reviews [12]. The current study protocol was prospectively registered in the International Prospective Register of Systematic Reviews (PROSPERO CRD420251241667).

### Eligibility criteria

#### Inclusion criteria

- Articles reporting cardiovascular events and new-onset diabetes mellitus, hypertension, dyslipidemia, and other metabolic effects following hysterectomy.
- Full-text articles in the English language
- Randomized controlled trials (RCTs), case-control studies, cross-sectional studies, cohort studies, case series, and experimental studies.

#### Exclusion criteria

- Systematic reviews, meta-analyses, or book chapters.

### Data extraction

Two reviewers (DMCSJ & AJ) conducted the initial data extraction independently, and a third reviewer (SS) cross-checked it. Finally, the fourth reviewer (TDKMJ) verified the process for completeness. The reviewers discussed conflicts and resolved discrepancies. Key study characteristics were extracted and organized into predefined tables concerning outcome measures: diabetes mellitus, hypertension, dyslipidemia, and other metabolic effects.

### Risk of bias and quality assessment

The quality of cohort studies was assessed using the Newcastle-Ottawa Scale (NOS), focusing on selection, comparability, and outcome, with a maximum score of 9 [13]. A modified version of NOS rated the quality of cross-sectional studies, including the domains of representativeness of sample, sample size, non-respondents, ascertainment of exposure, confounding factors, outcome assessment, and statistical analysis, with a maximum score of 10 [13]. The risk of bias and quality assessment employed a similar approach, as described in ‘data extraction,’ with conflicts resolved through discussion.

### Data synthesis

We performed statistical analyses using Review Manager (RevMan) version 5.4.1 [14]. We performed separate meta-analyses using a generic inverse-variance model with random effects for each outcome, stratifying by study design (cross-sectional vs. cohort) and effect measure (adjusted odds ratio [aOR], adjusted hazard ratio [aHR], or relative risk [RR]). For each study, we calculated the log-transformed effect estimate and its standard error from the reported 95% confidence intervals and entered them into RevMan. We assessed heterogeneity using the I² statistic; I² values ≥ 50% were considered high heterogeneity. When a single study reported multiple exposure groups (e.g., hysterectomy alone vs. hysterectomy with bilateral oophorectomy) sharing the same reference group, we pooled these estimates internally before inclusion. We performed sensitivity analyses by excluding larger studies to assess their influence on pooled estimates. We assessed publication bias using funnel plot asymmetry. We excluded studies from quantitative synthesis if they reported no measure of association (aOR, aHR, or RR) with 95% confidence intervals, or if there was only one study per outcome and no comparable studies for pooling. We performed narrative synthesis for outcomes that did not meet meta-analysis criteria. All pooled estimates are presented as summary effect sizes with 95% confidence intervals, accompanied by forest plots. Results were considered statistically significant at p < 0.05.

## Results

### Study characteristics

Fig 1 shows the PRISMA 2020 flow diagram for study selection. After reviewing 18,337 studies from four databases, 20 studies met the inclusion criteria. The included studies comprised 11 cohort studies [3,15–24], 8 cross-sectional studies [1,2,25–30], and one mixed study [31] with both cohort and cross-sectional components. 18 studies [1–3,16–29,31] contributed to the meta-analysis, which quantitatively synthesized the association between hysterectomy and hypertension, diabetes, hyperlipidemia, obesity, and metabolic syndrome. The studies were published from 1995 to 2026, with sample sizes ranging from 679 to over 9 million participants, and most publications were from the United States. Tables 1-4 summarize the key study characteristics, including study name, year of publication, ethnicity, sample size, study design, follow-up duration, and main results.

**Fig 1.**
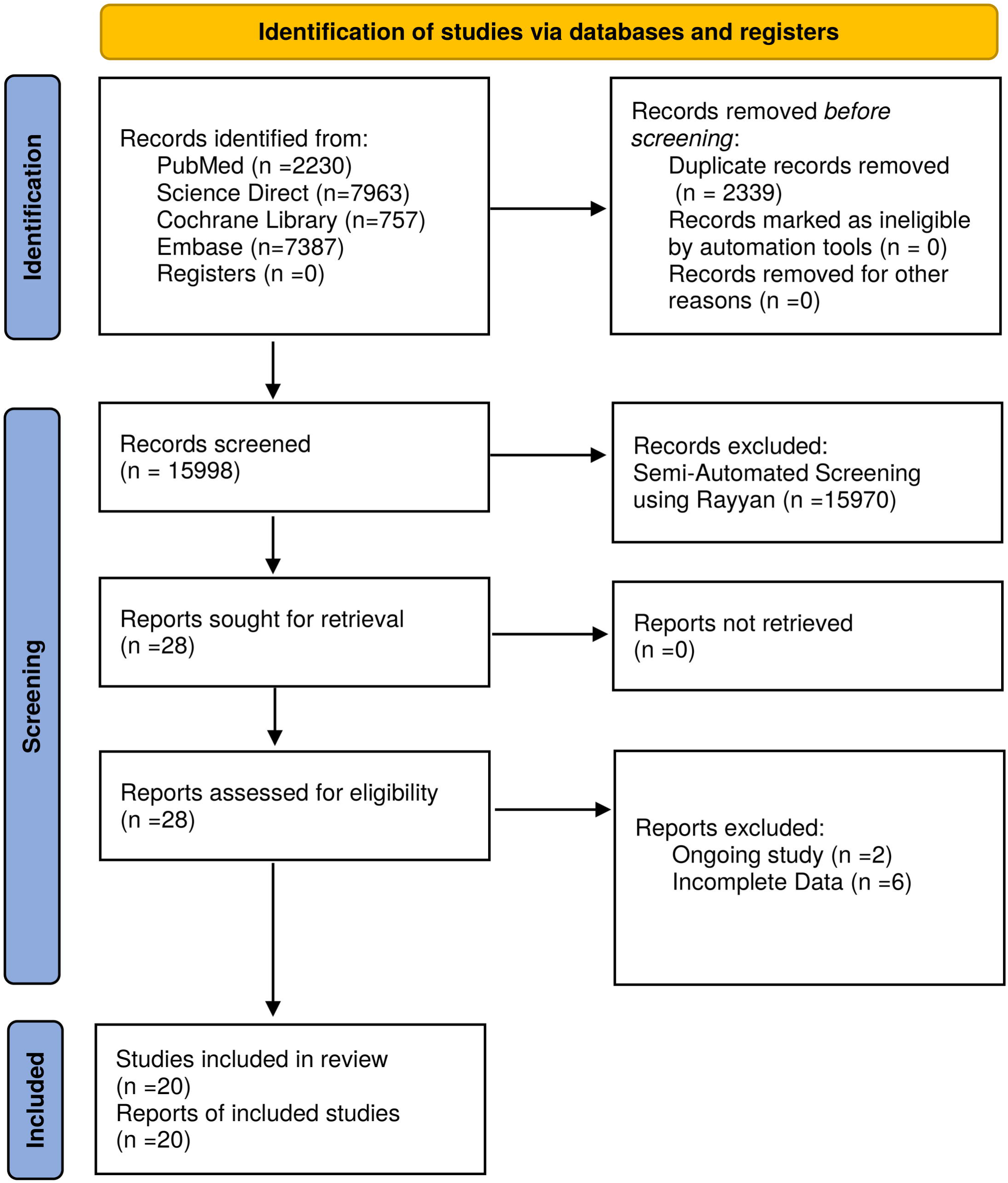
PRISMA 2020 flow diagram for study selection. Flow of records through identification, screening and inclusion, prepared according to the PRISMA 2020 statement.

**Table 1.** Characteristics of studies examining the association between hysterectomy and hypertension.

| Study Year | Population | Size (n) | Design | Follow-up (y) | Exposed | Unexposed |
|---|---|---|---|---|---|---|
| Luoto et al 1995 | Finnish women aged 30-95 (Mini-Finland Health Survey) | 3,780 | Cross-sectional | N/A | Hyst with ovarian preservation (n=163): aOR 2.18 (1.52-3.11): Mean DBP: 89.5 mmHg;<br>Hyst+BSO (n=55): aOR 1.98 (1.11-3.53): Mean DBP: 90.6 mmHg | n=3,562: OR 1.00; Mean DBP: 85.9 mmHg |
| Kharazmi et al 2007 | Finnish women aged 30-99 (Health 2000 Survey) | 2,514 | Cross-sectional | N/A | Hysterectomy (any) (n=591): aOR 1.24 (0.83-1.85); AF: 19.4% | n=1,923: OR 1.00 |
| Fletcher et al 2010 | Jamaican Black women | 679 | Cross-sectional | N/A | Hysterectomy (n=328): aOR 0.88 (0.62-1.23); Prevalence: 64%; Mean SBP 138 mmHg; Mean DBP 82 mmHg | n=341: OR 1.00; Prevalence: 69%; Mean SBP 144 mmHg; Mean DBP 86 mmHg |
| Matthews et al 2013 | US women (SWAN) | 1,952 | Prospective cohort | 11 | Hyst with ovarian preservation (n=77): Mean SBP 118.16 mmHg;<br>Hyst+BSO (n=106): Mean SBP 121.88 mmHg | n=1,769: Mean SBP 117.57 mmHg |
| Ding et al 2018 | Taiwanese women aged 30-49 (NHIRD) | 33,370 | Retrospective cohort | 6.4 | Hysterectomy (n=6,674): aHR 1.35 (1.27-1.44); 27.8/1000 PY | n=26,696: HR 1.00; 20.2/1000 PY |
| Laughlin-Tommaso et al 2018 | US women aged ≥18 (Minnesota -REP) | 4,188 | Prospective cohort (historical) | 21.9 | Hysterectomy with ovarian conservation (n=2,094): aHR 1.13 (1.03-1.25); 30-y risk: 62.6% | n=2,094: HR 1.00; 30-y risk: 56.1% |
| Halli et al 2020 | Indian women aged 15-49 (NFHS-4 2015-16) | 667,258 | Cross-sectional | N/A | Hysterectomy (n=640,709): aOR 1.72 (1.14-2.58); Age-adjusted prevalence: 11.9% | No hysterectomy (n=17,878): OR 1.00; Age-adjusted prevalence: 10.6% |
| Madika et al 2021 | French women aged 40-65 (E3N cohort) | Total: 50,286 | Prospective cohort | 16.4 | Hysterectomy (any) (n=8,511): aHR 1.18 (1.12-1.24); Incidence: 20/1000 PY | n=41,775: HR 1.00; Incidence: 17/1000 PY |
| Mustafa 2023 | Indian women aged ≥45 (LASI 2017-2018) | Total: 32,460 | Cross-sectional | N/A | Hysterectomy (n=3,390): aOR 1.36 (1.26-1.48); Prevalence: 42.5% | n=29,070: OR 1.00; Prevalence: 31.3% |
| Yan et al 2025 | US women (NHANES 2007-2018) | Total: 12,628 | Cross-sectional | N/A | Hysterectomy (n=2,858): aOR 1.26 (1.04-1.52) | n=9,770: OR 1.00 |
| Yan et al 2025 | European (UK Biobank/IEU Open GWAS) | GWAS: 462,933 total (46,411 cases, 416,522 controls) | Two-sample Mendelian randomization | N/A | Genetically predicted hysterectomy (28 SNPs) (n=46,411): OR 1.205 (1.043-1.392); Mean SBP 9.6 mmHg higher; Mean DBP 6.7mmHg higher | n=416,522: OR 1.00 |
| Yuk et al 2025 | Korean women aged 40-59 | 27,300 | Prospective cohort) | 10.6 | Hysterectomy (any) (n=13,650): aHR 1.11 (1.05-1.17); Incidence 22/1000 PY<br>Hysterectomy with ovarian preservation (n=13,515): aHR 1.15 (1.08-1.23) | n=13,650: HR 1.00; Incidence 20/1000 PY |
| Bi et al 2026 | Chinese women aged ≥18 (urban health screening) | 9,013,462 | Cross-sectional (multicenter) | N/A | Hysterectomy (n=210,793): aRR 1.22 (1.21-1.23) | n=8,802,669: RR 1.00 |

*Abbreviations: aHR, adjusted hazard ratio; aOR, adjusted odds ratio; aRR, adjusted relative risk; AF, attributable fraction; BSO, bilateral salpingo-oophorectomy; CI, confidence interval; DBP, diastolic blood pressure; Hb, hemoglobin; HR, hazard ratio; Hyst, hysterectomy; N/A, not applicable; NR, not reported; OR, odds ratio; AF, attributable fraction; PAF, population attributable fraction; PY, person-years; RR, relative risk; SBP, systolic blood pressure; SNPs, single nucleotide polymorphisms; PY, person years*

**Table 2.** Characteristics of studies examining the association between hysterectomy and diabetes.

| Study | Outcome | Population | Size | Design | Follow-up (y) | Exposed | Unexposed |
|---|---|---|---|---|---|---|---|
| Bi et al<br>2026 | DM | Chinese (urban) | n = 9,013,462 | Cross-sectional (multicenter) | N/A | Hysterectomy (n=210,793): RR 1.26 (1.24-1.28); Diabetes prevalence: 9.88% | n = 8,802,669<br>RR 1.00 (reference); Diabetes prevalence: 3.38% |
| Ouyang et al<br>2025 | DM | UK (Biobank) | n = 127,514 | Prospective cohort | Median: 12.8 | Hysterectomy alone (n=7,004): 1.20 (1.09-1.32); Hysterectomy + BSO (n=6,254): 1.19 (1.08-1.32); BSO alone (n=643): 1.02 (0.72-1.45) | n = 113,613<br>HR 1.00 (reference) |
| Chiang et al<br>2020 | DM | Taiwanese (NHIRD) | n = 34,933 | Retrospective cohort | Median: 7.1 | Overall hysterectomy (n=7,088): aHR 1.37 (1.23-1.52); Hysterectomy + oophorectomy (n=712): 1.28 (0.93-1.76); Incidence: 9.12 per 1000 person-years | n = 27,845<br>aHR 1.00 (reference); Incidence: 6.78 per 1000 person-years |
| Halli et al<br>2020 | Blood glucose (>140 mg/dl) | Indian (NFHS-4) | n = 653,002 | Cross-sectional | N/A | Hysterectomy (n=17,787):<br>OR 1.15 (1.10-1.21);<br>Prevalence: 12.2% | n = 635,215<br>OR 1.00 (reference); Prevalence: 5.7% |
| Rocca et al<br>2016 | DM | American (Minnesota) | n = 3,306 | Prospective cohort (historical) | Median: 14.5 | Bilateral oophorectomy (n=1,653): HR 1.19 (1.06-1.33) | n = 1,653<br>HR 1.00 (reference) |
| Matthews et al<br>2013 | HOMA-IR | American (SWAN multi-ethnic) | n = 1,952 | Prospective cohort | 11 years | Hyst with ovarian conservation (n=77): HOMA-IR 0.32 (p=0.58); Hyst + BSO (n=106): HOMA-IR 0.29 (p=0.78) | n = 1,769<br>HOMA-IR at FMP 0.30 |
| Laughlin-Tommaso et al<br>2018 | DM | American (Minnesota) | n = 4,188 | Prospective cohort | Median: 21.9 | Hysterectomy with ovarian conservation (n=2,094): HR 1.09 (0.97-1.22); Absolute risk at 30 years: 49.6% (46.3-53.0) | n = 2,094<br>HR 1.00 (reference); Absolute risk at 30 years: 45.9% (42.6-49.4) |
| Luo et al<br>2017 | DM | American (WHI) | n = 67,130 | Prospective cohort | Mean: 13.4 | Hysterectomy alone (n=10,591): 1.15 (1.07-1.24); Hysterectomy + BSO (n=13,761): 1.16 (1.08-1.24); BSO alone (no hysterectomy) (n=243): 1.20 (0.82-1.75); Incidence: 9.52 per 1000 person-years | n = 42,778<br>HR 1.00 (reference); Incidence: 7.55 per 1000 person-years |
| Appiah et al<br>2014 | DM | American (NHANES I/NHEFS) | n = 2,597 | Prospective cohort | Mean: 8.7 | Hysterectomy + BSO (n=484): 1.57 (1.03-2.41): Incidence: 8.5 per 1000 person-years; Hysterectomy alone (n=551): 1.38 (0.94-2.04):Incidence: 8.2 per 1000 person-years;<br>Any hysterectomy: 1.46 (1.04-2.04) | n = 1,562<br>HR 1.00 (reference); Incidence: 7.4 per 1000 person-years |
| Fletcher et al<br>2010 | DM | Jamaican (Black) | n = 679 | Cross-sectional (observational) | N/A | n = 328 (hysterectomy)<br>Diabetes prevalence 20%; Fasting glucose 5.3 mmol/L (5.2-5.5); Adjusted OR 1.31 (0.86-1.97) | n = 341<br>Diabetes prevalence 18%; Fasting glucose 5.2 mmol/L (5.1-5.4); OR 1.00 (reference) |
| Luoto et al<br>1995 | DM | Finnish | n = 3,780 | Cross-sectional (population-based) | N/A | n = 163 (hysterectomy with ovarian preservation)<br>Mean blood glucose 5.4 mmol/L (5.2-5.6); Diabetes prevalence: 5.5% | n = 3,562<br>Mean blood glucose 5.4 mmol/L (5.4-5.5); Diabetes prevalence: 6.4% |

*Abbreviations: aHR, adjusted hazard ratio; BSO, bilateral salpingo-oophorectomy; CI, confidence interval; FMP, final menstrual period; HOMA-IR, homeostasis model assessment-insulin resistance; HR, hazard ratio; IQR, interquartile range; NHANES, National Health and Nutrition Examination Survey; NHEFS, NHANES I Epidemiologic Follow-up Study; NHIRD, National Health Insurance Research Database; NFHS-4, National Family Health Survey round 4; NR, not reported; OR, odds ratio; RR, relative risk; SWAN, Study of Women's Health Across the Nation; UK, United Kingdom; WHI, Women's Health Initiative.*

**Table 3.** Characteristics of studies examining the association between hysterectomy and hyperlipidemia.

| Study | Outcome | Population | Design | Follow-up (y) | Exposed | Unexposed |
|---|---|---|---|---|---|---|
| Luoto et al 1995 | Lipid profile | 3,780 Finnish | Cross-sectional | N/A | Hyst with ovarian preservation (n=163): TC 7.1 mmol/L; HDL 1.8 mmol/L; TG 1.5 mmol/L; BMI 26.9 kgm<sup>-2</sup><br>Hyst+BSO (n=55): TC 7.6 mmol/L; HDL 1.8 mmol/L; TG 1.5 mmol/l; BMI 27.3 kgm<sup>-2</sup> | n=3,562: TC 7.0 mmol/L; HDL 1.8 mmol/L; TG 1.4 mmol/L; BMI 25.9 kgm<sup>-2</sup> |
| Zhang et al 2005 | Lipid profile | 1,726 American Indian (Strong Heart Study) | Cross-sectional | N/A | Hyst with ovarian preservation (n=161): TC 200.9 mg/dl; LDL 125.4 mg/dl; HDL 48.9 mg/dl<br>Hyst + oophorectomy (n=238): TC 193.9mg/dl; LDL 120.4 mg/dl; HDL 46 mg/dl | n=1,327: TC 193.3 mg/dl; LDL 118.2; HDL 47.4 mg/dl |
| Fletcher et al 2010 | Lipid profile | 679 Jamaican Black | Cross-sectional | N/A | Hysterectomy (n=328): TC 6.0 mmol/L; HDL 1.3 mmol/L; TG 1.0 mmol/L | n=341: TC 5.7 mmol/L; HDL 1.3 mmol/L; TG 0.96 mmol/L |
| Matthews et al 2013 | Lipid profile | 1,952 American (SWAN) | Prospective cohort | Up to 11 | Hyst with ovarian preservation (n=77): LDL 122.08 mg/dl; HDL 55.22 mg/dl; TG 111.05 mg/dl; ApoB 115.87 mg/dl<br>Hyst+BSO (n=106): LDL 114.79 mg/dl; HDL 54.78 mg dL; TG 111.41 mg/dl; ApoB 108.54 mg/dl | n=1,769: LDL 115.19 mg/dl; HDL 57.52 mg/dl; TG 109.03 mg/dl; ApoB 110.2 mg/dl |
| Rocca et al 2016 | Hyperlipidemia | 3,306 American (Minnesota) | Prospective cohort (historical) | Median 14.5 | BSO before 50 (n=1,653): aHR 1.19 (1.07-1.32); ≤45 y: aHR 1.23 (1.06-1.42); Age 46-49: aHR 1.19 (1.02-1.38) | Referent women (n=1,653): HR 1.00 |
| Laughlin-Tommaso et al 2018 | Hyperlipidemia | 4,188 American (Minnesota) | Prospective cohort | Median 21.9 | Hyst with ovarian conservation (n=2,094): aHR 1.14 (1.05-1.25); 30-y risk: 73.1% | n=2,094: HR 1.00; 30-y risk: 69.3% |
| Li et al 2018 | Hyperlipidemia | 35,030 Taiwanese (NHIRD) | Retrospective cohort | Median 6.0 | Any hyst/ooph (n=7,006): aHR 1.27 (1.19-1.35); Hyst only (n=5,887): aHR 1.27 (1.19-1.35); Hyst+BSO (n=563): aHR 1.51 (1.31-1.75); Ooph only (n=556): aHR 1.11 (0.93-1.32); Incidence: Hyst only 3.43/100 PY; Hyst+BSO 4.93/100 PY; Ooph only 3.51/100 PY | n=28,024: HR 1.00; Incidence: 2.65/100 PY |
| Bi et al | Dyslipidemia | 9,013,462 Chinese (urban) | Cross-sectional (multicenter) | N/A | Hysterectomy (n=210,793): RR 1.18 (1.16-1.19) | n=8,802,669: RR 1.00 |

*Abbreviations: aHR, adjusted hazard ratio; aOR, adjusted odds ratio; ApoB, apolipoprotein B; BMI, body mass index; BSO, bilateral salpingo-oophorectomy; CI, confidence interval; HDL, high-density lipoprotein; HR, hazard ratio; Hyst, hysterectomy; TC, Total Cholesterol; TG, Triglyceride;  LDL, low-density lipoprotein; MAFLD, metabolic dysfunction-associated fatty liver disease; MESA, Multi-Ethnic Study of Atherosclerosis; N/A, not applicable; NHIRD, National Health Insurance Research Database; NR, not reported; Ooph, oophorectomy; OR, odds ratio; PY, person-years; RR, relative risk; SWAN, Study of Women's Health Across the Nation*

**Table 4.** Characteristics of studies examining the association between hysterectomy and obesity or metabolic syndrome.

| Study | Outcome | Population | Size | Design | Follow-up (y) | Exposed | Unexposed |
|---|---|---|---|---|---|---|---|
| Luoto et al 1995 | BMI | Finnish | 3,780 | Cross-sectional | N/A | Hyst with ovarian preservation (n=163): Mean 26.9 kgm<sup>-2</sup>; Hyst+BSO (n=55): Mean 27.3 kgm<sup>-2</sup> | No hysterectomy (n=3,562): Mean 25.9 kgm<sup>-2</sup> |
| Fletcher et al 2010 | Obesity (BMI ≥30) | Jamaican Black | 679 | Cross-sectional | N/A | Hysterectomy (n=328): aOR 1.04 (0.74-1.45) | No hysterectomy (n=341): OR 1.00 |
| Laughlin-Tommaso et al 2018 | Obesity | American (Minnesota) | 4,188 | Prospective cohort | Median 21.9 | Hyst with ovarian conservation (n=2,094): aHR 1.18 (1.04-1.35); 30-y risk: 36.0% | Referent women (n=2,094): HR 1.00; 30-y risk: 31.7% |
| Broni et al 2024 | Metabolic syndrome (prevalent) | Multi-ethnic US (MESA) | 3,367 | Cross-sectional | N/A | Hyst only (n=404): aOR 1.52 (1.17-1.98); Hyst+BSO (n=723): aOR 1.38 (1.20-1.72); Ooph only (n=91): aOR 1.32 (0.77-2.28) | No hyst/no ooph (n=2,149): OR 1.00 |
| Broni et al 2024 | Metabolic syndrome (incident) | Multi-ethnic US (MESA) | 1,355 | Prospective cohort | Median 10.5 | Hyst only (n=397): aHR 1.32 (1.01-1.73); Hyst+BSO (n=NR): aHR 1.40 (1.13-1.74); Ooph only (n=303): aHR 0.63 (0.34-1.15) | No hyst/no ooph (n=958): HR 1.00 |
| Bi et al 2026 | Obesity | Chinese (urban) | 9,013,462 | Cross-sectional (multicenter) | N/A | Hysterectomy (n=210,793): RR 1.31 (1.29-1.32); Prevalence: 16.6% obese | No hysterectomy (n=8,802,669): RR 1.00; Prevalence: 8.48% obese |
| Bi et al 2026 | MAFLD | Chinese (urban) | 9,013,462 | Cross-sectional (multicenter) | N/A | Hysterectomy (n=210,793): RR 1.25 (1.24-1.26) | No hysterectomy (n=8,802,669): RR 1.00 |
| Bi et al 2026 | Metabolic syndrome | Chinese (urban) | 9,013,462 | Cross-sectional (multicenter) | N/A | Hysterectomy (n=210,793): RR 1.18 (1.16-1.21) | No hysterectomy (n=8,802,669): RR 1.00 |

*Abbreviations: aHR, adjusted hazard ratio; aOR, adjusted odds ratio; BMI, body mass index; BSO, bilateral salpingo-oophorectomy; CI, confidence interval; HR, hazard ratio; MAFLD, metabolic dysfunction-associated fatty liver disease; MESA, Multi-Ethnic Study of Atherosclerosis; N/A, not applicable; NR, not reported; Ooph, oophorectomy; OR, odds ratio; PY, person-years; RR, relative risk*

### Risk of bias and quality assessment

The Newcastle-Ottawa Scale (NOS) assessed the cohort studies and rated all as high quality. Six studies achieved the maximum score of 9 [3,15,19–21,23], while five studies scored 8 out of 9 [16–18,22,31]. Chiang et al. received a score of 7 out of 9, mainly because the exposed cohort was not representative and comparability was a concern [24]. The modified Newcastle-Ottawa Scale assessed the cross-sectional studies and rated the studies as average to good quality, with scores ranging from 6 to 9 out of 10. Bi et al. received the highest score of 9 [29], while two studies received 8 [30,31] and four received 7 [1,25,26,28]. Two studies received scores of 6, primarily due to limitations in sample size justification and representativeness [2,27]. A detailed quality assessment of the studies is shown in Tables 5 and 6. Visual assessment of the publication bias revealed minimal asymmetry in the funnel plot. Egger’s test was not performed because of the limited number of studies for each outcome.

**Table 5.** Quality assessment of cohort studies using the Newcastle-Ottawa Scale.

|  | Selection |  |  |  | Comparability | Outcome |  |  |  |
|---|---|---|---|---|---|---|---|---|---|
| **Study, year (ref)** | **Representativeness of the exposed cohort** | **Selection of the non-exposed cohort** | **Ascertainment of exposure** | **Demonstration that outcome of interest was not present at start of study** | **Comparability of cohorts on the basis of the design or analysis** | **Assessment of outcome** | **Was follow-up long enough for outcomes to occur** | **Adequacy of follow-up of cohorts** | **Total** |
| Matthews 2013 | 1 | 1 | 1 | 1 | 2 | 1 | 1 | 1 | 9/9 |
| Ding 2018 | 1 | 1 | 1 | 1 | 2 | 1 | 1 | 0 | 8/9 |
| Laughlin-Tommaso 2018 | 0 | 1 | 1 | 1 | 2 | 1 | 1 | 1 | 8/9 |
| Madika 2021 | 1 | 1 | 1 | 1 | 2 | 1 | 1 | 1 | 9/9 |
| Yuk 2025 | 1 | 1 | 1 | 1 | 2 | 1 | 1 | 1 | 9/9 |
| Rocca 2016 | 1 | 1 | 1 | 1 | 2 | 1 | 1 | 1 | 9/9 |
| Li 2018 | 1 | 1 | 1 | 1 | 2 | 1 | 1 | 0 | 8/9 |
| Ouyang 2025 | 1 | 1 | 1 | 1 | 2 | 1 | 1 | 1 | 9/9 |
| Chiang 2020 | 0 | 1 | 1 | 1 | 1 | 1 | 1 | 1 | 7/9 |
| Luo 2017 | 1 | 1 | 1 | 1 | 2 | 1 | 1 | 1 | 9/9 |
| Appiah 2014 | 1 | 1 | 1 | 1 | 2 | 0 | 1 | 1 | 8/9 |
| Broni 2024 | 1 | 1 | 1 | 0 | 2 | 1 | 1 | 1 | 8/9 |

*Newcastle-Ottawa Scale domains are scored as follows: Selection (maximum 4 stars), Comparability (maximum 2 stars), Outcome (maximum 3 stars); higher scores indicate lower risk of bias.*

**Table 6.** Quality assessment of cross-sectional studies using the modified Newcastle-Ottawa Scale.

| First author & reference | Selection |  |  |  | Comparability | Outcome |  | Score |
|---|---|---|---|---|---|---|---|---|
|  | **Representativeness of the sample** | **Sample size justified** | **Non-respondents** | **Ascertainment of exposure (max 2)** | **Confounding controlled (max 2)** | **Outcome assessment (max 2)** | **Statistics** | **Total** |
| Luoto 1995 | 1 | 0 | 0 | 2 | 2 | 1 | 1 | 7/10 |
| Kharazmi 2007 | 1 | 0 | 1 | 1 | 2 | 1 | 1 | 7/10 |
| Fletcher 2010 | 0 | 0 | 0 | 2 | 2 | 1 | 1 | 6/10 |
| Halli 2020 | 1 | 0 | 0 | 2 | 2 | 1 | 1 | 7/10 |
| Mustafa 2023 | 1 | 1 | 0 | 1 | 2 | 1 | 1 | 7/10 |
| Yan 2025 | 1 | 0 | 0 | 1 | 2 | 1 | 1 | 6/10 |
| Bi 2026 | 1 | 1 | 0 | 2 | 2 | 2 | 1 | 9/10 |
| Zhang 2005 | 1 | 0 | 0 | 2 | 2 | 2 | 1 | 8/10 |
| Broni 2024 | 1 | 0 | 0 | 2 | 2 | 2 | 1 | 8/10 |

*Modified Newcastle-Ottawa Scale domains are scored as follows: Selection (maximum 5 points), Comparability (maximum 2 points), Outcome (maximum 3 points); higher scores indicate lower risk of bias.*

### Synthesis of results

#### Sensitivity analysis

The largest study by Bi et al included over 9 million Chinese women [29]. In the meta-analysis of cross-sectional studies exploring the association between hysterectomy and hypertension, excluding Bi 2026 resulted in a pooled adjusted odds ratio of 1.37 (95% CI: 1.15 to 1.64) with I² of 69%, compared to the original pooled estimate of 1.32 (95% CI: 1.18 to 1.48), indicating a minimal effect on the overall estimate. However, in the meta-analysis focusing on the association between hysterectomy and diabetes, excluding Bi 2026 attenuated the pooled estimate from 1.21 (95% CI: 1.11 to 1.32) to 1.15 (95% CI: 1.10 to 1.21). Heterogeneity was eliminated (I² = 0%), suggesting that the large Chinese study was the primary source of heterogeneity. Similarly, in the meta-analysis of obesity and hysterectomy, excluding Bi 2026 left only Fletcher et al. [27], which had an odds ratio of 1.04 (95% CI: 0.74 to 1.46), showing that the significant pooled estimate of 1.24 depended entirely on the inclusion of the large Chinese study. Furthermore, sensitivity analyses were applied systematically across all other outcomes using leave-one-out analyses. However, excluding any single study did not substantially change the results.

### Association between hysterectomy and hypertension

#### Evidence from cross-sectional studies

Seven cross-sectional studies [1,2,25–29] with a total of 10,183,086 participants showed that hysterectomy was associated with a 32% increase in the odds of hypertension, with a pooled adjusted odds ratio of 1.32 and a 95% confidence interval ranging from 1.18 to 1.48. The test for overall effect was highly significant (Z = 4.74, P < 0.00001). Heterogeneity was substantial, with an I² statistic of 76% (P = 0.0003). Fig 2 shows the forest plot for the meta-analysis of cross-sectional studies.

**Fig 2.**
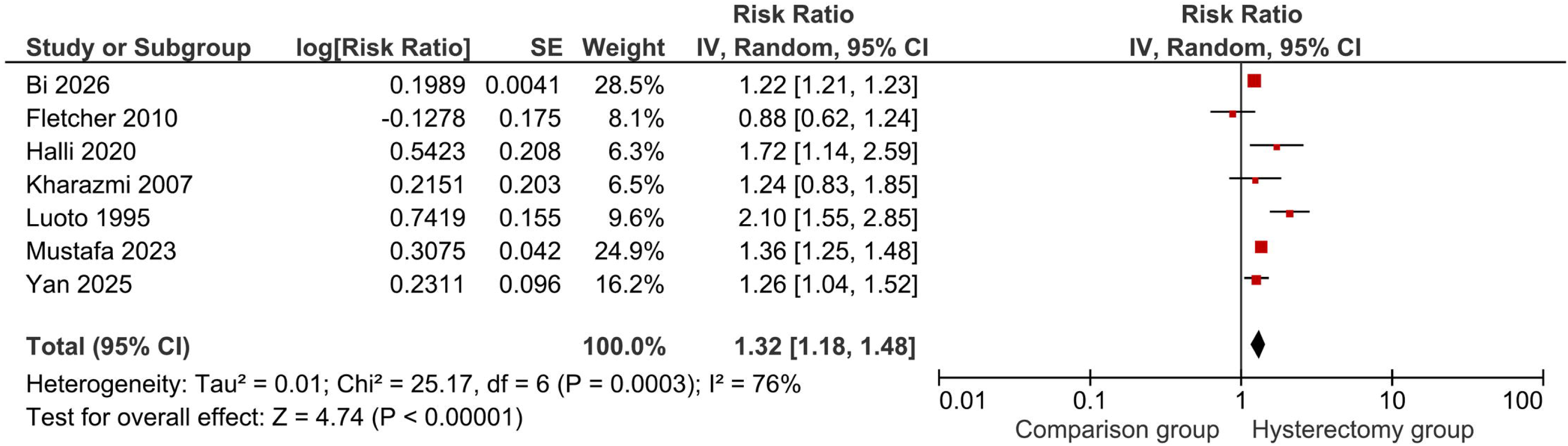
Association between hysterectomy and hypertension (cross-sectional studies). Squares show study-specific effect estimates (square size is proportional to the study weight in the random-effects model) and horizontal lines show 95% confidence intervals. The diamond shows the pooled estimate, with its width representing the 95% confidence interval. Effect estimates are plotted on a logarithmic scale; values above 1 indicate higher risk in the hysterectomy group than in the comparison group. Estimates were pooled using an inverse-variance random-effects model. Pooled effect measure: risk ratio. Heterogeneity is summarized by Tau², Chi² and I². CI, confidence interval; IV, inverse variance; SE, standard error.

#### Evidence from cohort studies

Four cohort studies [17–20] with a total of 111,374 participants demonstrated that hysterectomy was associated with a 19% increased risk of developing hypertension, with a pooled adjusted hazard ratio of 1.19 and a 95% confidence interval of 1.09 to 1.30. The test for overall effect was highly significant (Z = 3.92, P < 0.0001). Heterogeneity was considerable, with an I² statistic of 87% (P < 0.0001). Fig 3 shows the forest plot for the cohort study meta-analysis.

**Fig 3.**
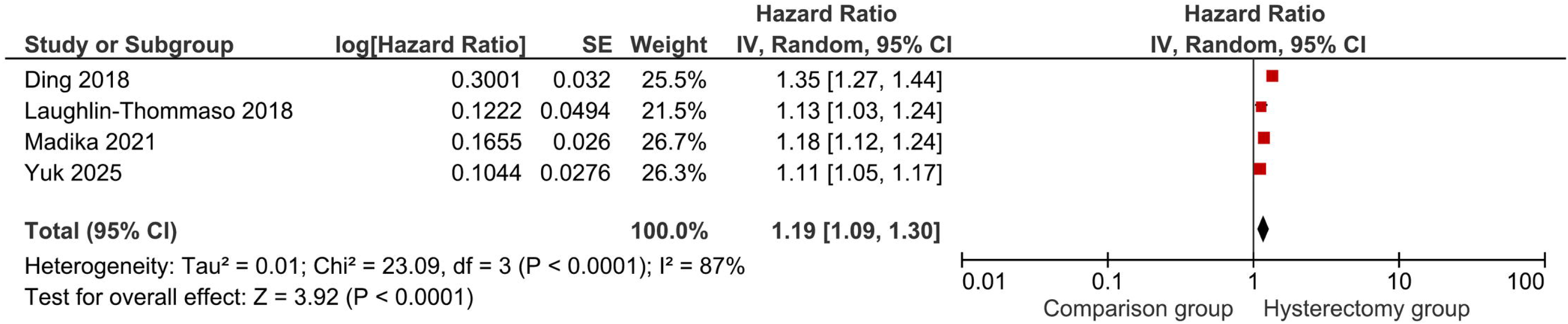
Association between hysterectomy and hypertension (cohort studies). Squares show study-specific effect estimates (square size is proportional to the study weight in the random-effects model) and horizontal lines show 95% confidence intervals. The diamond shows the pooled estimate, with its width representing the 95% confidence interval. Effect estimates are plotted on a logarithmic scale; values above 1 indicate higher risk in the hysterectomy group than in the comparison group. Estimates were pooled using an inverse-variance random-effects model. Pooled effect measure: hazard ratio. Heterogeneity is summarized by Tau², Chi² and I². CI, confidence interval; IV, inverse variance; SE, standard error.

The prospective cohort study by Matthews et al., reporting the association between hysterectomy and hypertension, was excluded from the meta-analysis due to differences in outcome reporting [15]. The study followed up 1,952 American women for 11 years and reported mean systolic blood pressure values. Women with hysterectomy ± oophorectomy (n = 183) had higher mean systolic blood pressures (118.16 – 121.88 mmHg) compared to women without hysterectomy (117.57 mmHg) (n = 1769) [15].

#### Evidence linked to genetics

Yan et al., in their Mendelian randomization study, analyzed genetic variants associated with hysterectomy (28 single-nucleotide polymorphisms) in European women from the UK Biobank and the IEU Open GWAS database, comprising 46,411 cases and 416,522 controls [2]. Genetically predicted hysterectomy was associated with an odds ratio of 1.205 (95% CI: 1.043 to 1.392) for hypertension. In addition, genetically predicted hysterectomy was associated with higher mean systolic (9.6 mmHg) and diastolic (6.7 mmHg) blood pressure compared with unexposed women.

### Association between hysterectomy and diabetes

#### Evidence from cross-sectional studies

Three cross-sectional studies with a total of 9,667,143 participants [1,27,29] reported that hysterectomy was associated with a 21% increase in the odds of diabetes, with a pooled estimate of 1.21 and a 95% confidence interval of 1.11 to 1.32. The test for overall effect was highly significant (Z = 4.49, P < 0.00001). Heterogeneity was high, with an I² statistic of 84% (P = 0.002). Fig 4 shows the forest plot for the meta-analysis of cross-sectional studies.

**Fig 4.**
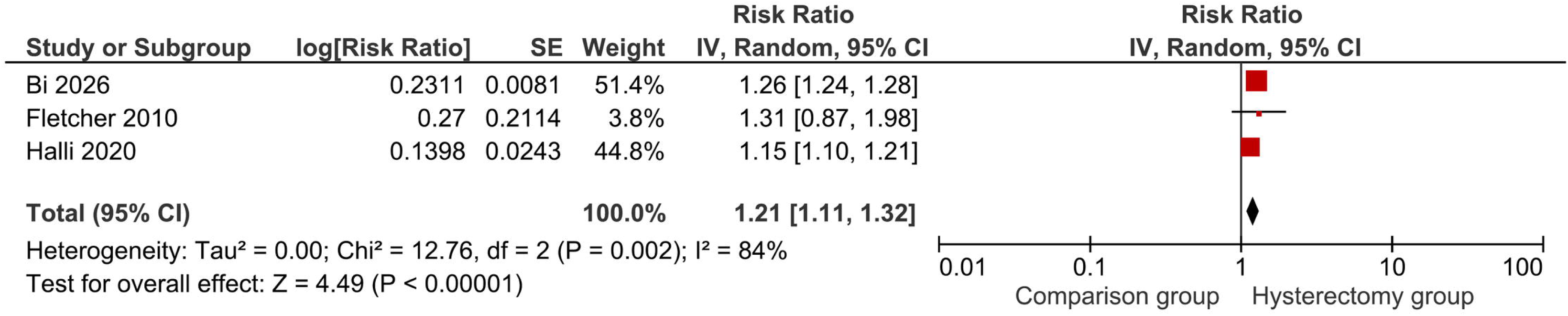
Association between hysterectomy and diabetes (cross-sectional studies). Squares show study-specific effect estimates (square size is proportional to the study weight in the random-effects model) and horizontal lines show 95% confidence intervals. The diamond shows the pooled estimate, with its width representing the 95% confidence interval. Effect estimates are plotted on a logarithmic scale; values above 1 indicate higher risk in the hysterectomy group than in the comparison group. Estimates were pooled using an inverse-variance random-effects model. Pooled effect measure: risk ratio. Heterogeneity is summarized by Tau², Chi² and I². CI, confidence interval; IV, inverse variance; SE, standard error.

#### Evidence from cohort studies

Six prospective cohort studies with a total of 239,650 participants [3,16,18,21,23,24] showed that hysterectomy was associated with a 20% increased risk of diabetes, with a pooled adjusted hazard ratio of 1.20 and a 95% confidence interval of 1.13 to 1.29. The test for overall effect was highly significant (Z = 5.38, P < 0.00001). Heterogeneity was moderate, with an I² statistic of 54% (P = 0.05). Fig 5 shows the forest plot for the meta-analysis of cohort studies.

**Fig 5.**
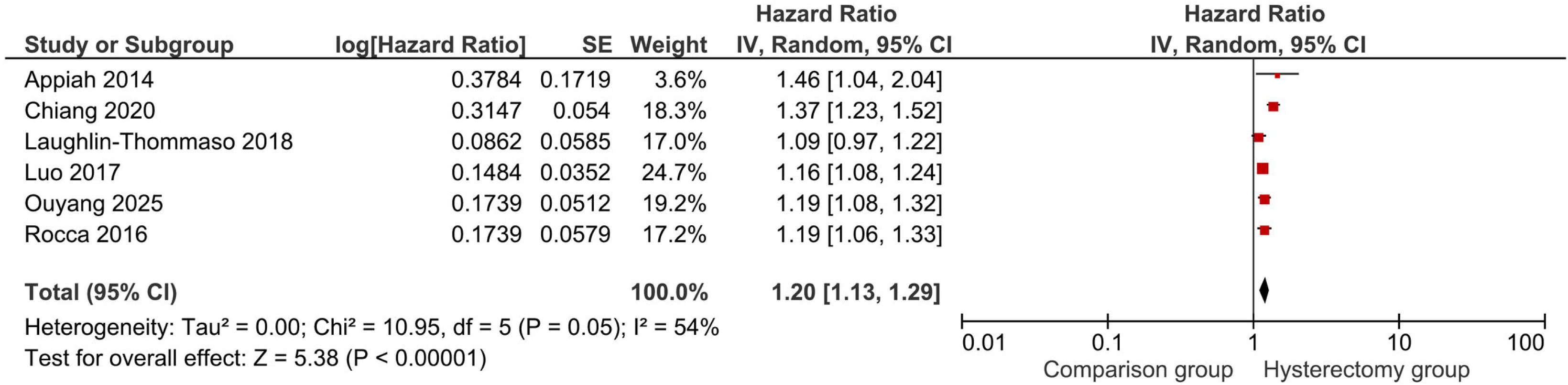
Association between hysterectomy and diabetes (cohort studies). Squares show study-specific effect estimates (square size is proportional to the study weight in the random-effects model) and horizontal lines show 95% confidence intervals. The diamond shows the pooled estimate, with its width representing the 95% confidence interval. Effect estimates are plotted on a logarithmic scale; values above 1 indicate higher risk in the hysterectomy group than in the comparison group. Estimates were pooled using an inverse-variance random-effects model. Pooled effect measure: hazard ratio. Heterogeneity is summarized by Tau², Chi² and I². CI, confidence interval; IV, inverse variance; SE, standard error.

### Association between hysterectomy and changes in lipid profile

#### Evidence on hyperlipidemia/dyslipidemia

Three cohort studies with a total of 42,524 participants [18,21,22] found that hysterectomy was associated with a 21% increased risk of hyperlipidemia, with a pooled adjusted hazard ratio of 1.21 and a 95% confidence interval of 1.13 to 1.29. The test for overall effect was highly significant (Z = 5.34, P < 0.00001). Heterogeneity was moderate, with an I² statistic of 51% (P = 0.13). Fig 6 shows the forest plot for the meta-analysis of cohort studies.

**Fig 6.**
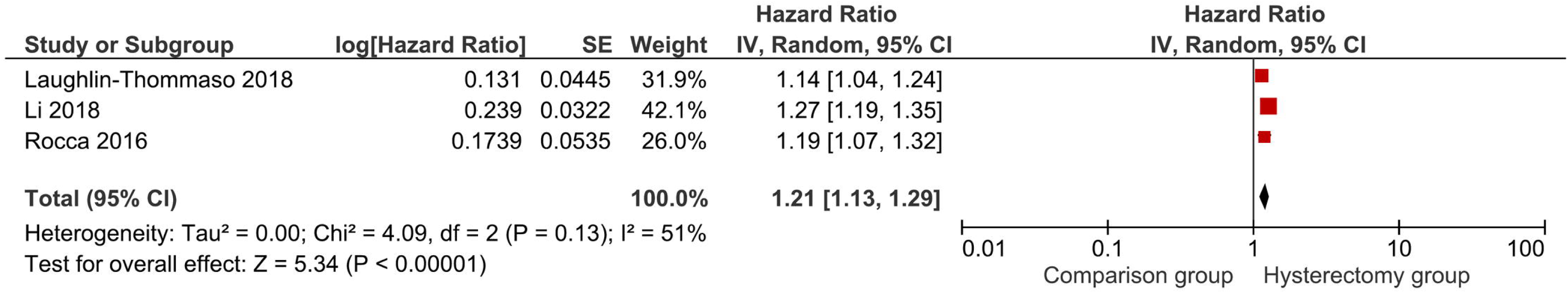
Association between hysterectomy and hyperlipidemia (cohort studies). Squares show study-specific effect estimates (square size is proportional to the study weight in the random-effects model) and horizontal lines show 95% confidence intervals. The diamond shows the pooled estimate, with its width representing the 95% confidence interval. Effect estimates are plotted on a logarithmic scale; values above 1 indicate higher risk in the hysterectomy group than in the comparison group. Estimates were pooled using an inverse-variance random-effects model. Pooled effect measure: hazard ratio. Heterogeneity is summarized by Tau², Chi² and I². CI, confidence interval; IV, inverse variance; SE, standard error.

In a cross-sectional study of over 9 million Chinese women, Bi et al. reported that hysterectomy was associated with a relative risk of 1.18 (95% CI: 1.16 to 1.19) of developing dyslipidemia [29]. No other cross-sectional studies reported dyslipidemia with comparable effect measures. Therefore, pooling was not possible for this outcome.

#### Evidence on mean lipid values

Several cross-sectional and cohort studies reported only mean lipid values without standard deviations or comparable effect measures, precluding meta-analysis. Luoto et al. showed that Finnish women who underwent hysterectomy (n = 218) had a mean total cholesterol of 7.22 mmol/L, compared with 7.0 mmol/L in unexposed women (n = 3562) [25]. High-density lipoprotein was similar across both groups (1.8 mmol/L). Triglycerides were 1.5 mmol/L in the hysterectomy group compared to 1.4 mmol/L in unexposed women. Zhang et al. studied American Indian women and reported that those who had undergone hysterectomy (n = 399) had a mean total cholesterol of 196.7 mg/dL, a mean low-density lipoprotein of 122.4 mg/dL, and a mean high-density lipoprotein of 47.2 mg/dL [30]. In comparison, unexposed women (n = 1327) had a mean total cholesterol of 193.3 mg/dL, a mean low-density lipoprotein of 118.2 mg/dL, and a mean high-density lipoprotein of 47.4 mg/dL [30]. Fletcher et al. found that Jamaican women who had hysterectomy (n = 328) had a mean total cholesterol of 6.0 mmol/L, a mean high-density lipoprotein of 1.3 mmol/L, and mean triglycerides of 1.0 mmol/L, compared with 5.7 mmol/L, 1.3 mmol/L, and 0.96 mmol/L, respectively, in unexposed women [27]. Matthews et al., in a prospective cohort study, reported that women with hysterectomy (n = 183) had a mean low-density lipoprotein of 117.85 mg/dL, a mean high-density lipoprotein of 54.96 mg/dL, a mean triglyceride of 111.25 mg/dL, and a mean apolipoprotein B of 111.62 mg/dL [15]. Unexposed women (n = 1769) had a mean low-density lipoprotein of 115.19 mg/dL, a mean high-density lipoprotein of 57.52 mg/dL, a mean triglyceride of 109.03 mg/dL, and a mean apolipoprotein B of 110.2 mg/dL [15].

### Evidence on obesity

Two cross-sectional studies [27,29] with a total of 9,014,141 participants found that hysterectomy was associated with a 24% increase in the odds of obesity, with a pooled estimate of 1.24 and a 95% confidence interval of 1.03 to 1.50. The test for overall effect was significant (Z = 2.27, p = 0.02), along with minimal heterogeneity (I^2^ = 45%, P = 0.18). Fig 7 shows the forest plot for the meta-analysis of cohort studies.

**Fig 7.**
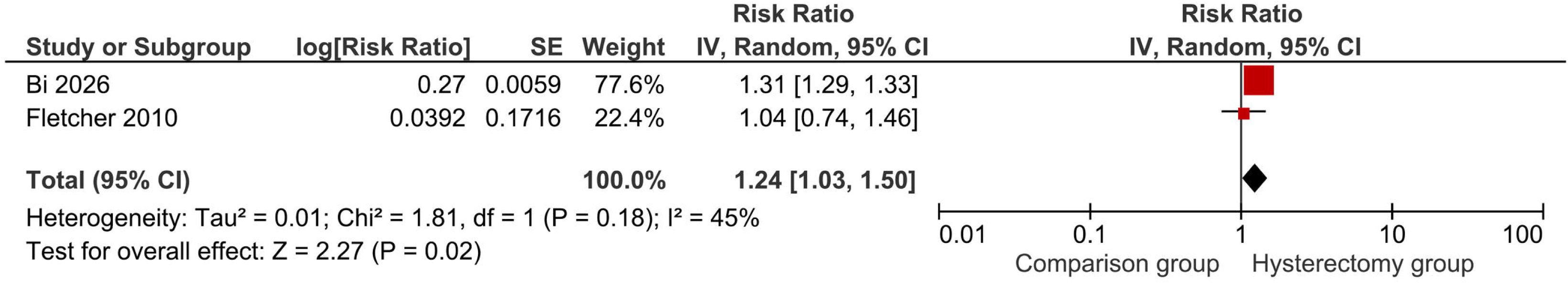
Association between hysterectomy and obesity (cross-sectional studies). Squares show study-specific effect estimates (square size is proportional to the study weight in the random-effects model) and horizontal lines show 95% confidence intervals. The diamond shows the pooled estimate, with its width representing the 95% confidence interval. Effect estimates are plotted on a logarithmic scale; values above 1 indicate higher risk in the hysterectomy group than in the comparison group. Estimates were pooled using an inverse-variance random-effects model. Pooled effect measure: risk ratio. Heterogeneity is summarized by Tau², Chi² and I². CI, confidence interval; IV, inverse variance; SE, standard error.

In the cohort study by Laughlin-Tommaso et al., 4,188 American women (Hysterectomy group – 2094 and comparison group – 2094) were followed for a median of 21.9 years, revealing an 18% increase in obesity in women with hysterectomy. They reported an adjusted hazard ratio of 1.18 (95% CI, 1.04 to 1.35). The 30-year risk of obesity was 36% among women who underwent hysterectomy compared with 31.7% among unexposed women [18]. Luoto et al. reported mean body mass index (BMI) values without standard deviations. Women with hysterectomy (n = 218) had a mean body mass index of 27 kg/m², and unexposed women (n = 3562) had a mean of 25.9 kg/m² [25].

### Evidence on metabolic syndrome

Two cross-sectional studies [29,31] with a total of 9,014,817 participants revealed that hysterectomy was associated with a 24% increase in the odds of metabolic syndrome, with a pooled estimate of 1.24 and a 95% confidence interval ranging from 1.08 to 1.44. The test for overall effect was highly significant (Z = 2.95, P = 0.003). However, the heterogeneity was high (I^2^ = 77%, P = 0.09). Fig 8 shows the forest plot for the meta-analysis of cross-sectional studies.

**Fig 8.**
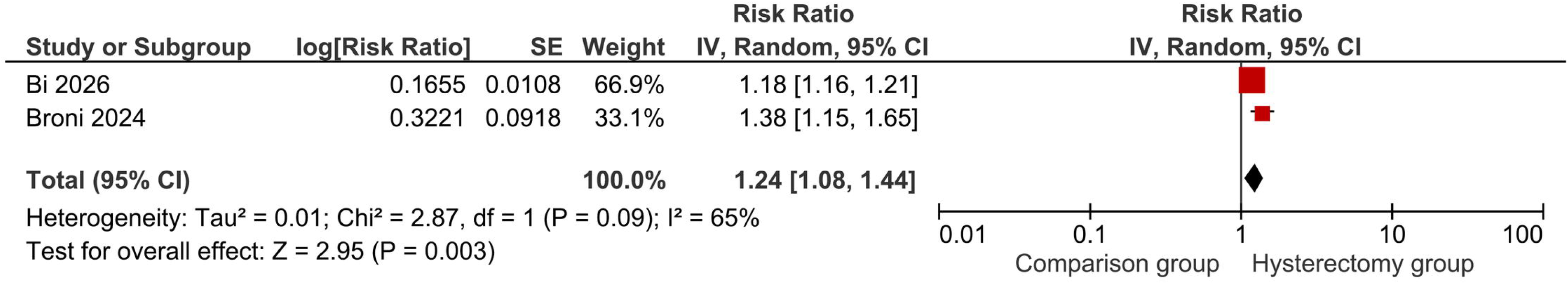
Association between hysterectomy and metabolic syndrome (cross-sectional studies). Squares show study-specific effect estimates (square size is proportional to the study weight in the random-effects model) and horizontal lines show 95% confidence intervals. The diamond shows the pooled estimate, with its width representing the 95% confidence interval. Effect estimates are plotted on a logarithmic scale; values above 1 indicate higher risk in the hysterectomy group than in the comparison group. Estimates were pooled using an inverse-variance random-effects model. Pooled effect measure: risk ratio. Heterogeneity is summarized by Tau², Chi² and I². CI, confidence interval; IV, inverse variance; SE, standard error.

The prospective cohort arm of Broni et al. followed 1,355 participants from the Multi-Ethnic Study of Atherosclerosis for a median of 10.5 years. They reported an adjusted hazard ratio of 1.32 (95% CI: 1.01 to 1.73) for metabolic syndrome in the hysterectomy group (n = 397) vs the unexposed group (n = 958) [31].

### Evidence on metabolic dysfunction-associated fatty liver disease (MAFLD)

Bi et al. reported that hysterectomy was associated with a 25% increased risk of MAFLD, based on a study of over 9 million urban Chinese women (Hysterectomy group – 210,793; comparison group – 8,802,669). They reported a relative risk of 1.25 (95% CI: 1.24 to 1.26) [29]. Table 7 summarizes the pooled estimates from meta-analyses of studies examining the association between hysterectomy and cardio-metabolic outcomes.

**Table 7.** Pooled estimates from meta-analyses examining the association between hysterectomy and cardio-metabolic outcomes.

| **Outcome** | **Study design** | **No. of studies** | **Total participants** | **Pooled estimate** | **95% CI** | **Test for overall effect** | **Heterogeneity (I<sup>2</sup>)** |
|---|---|---|---|---|---|---|---|
| Hypertension | Cross-sectional | 7 | 10,183,086 | OR 1.32 | 1.18–1.48 | Z = 4.74, P < .001 | 76% (P < .001) |
| Hypertension | Cohort | 4 | 111,374 | HR 1.19 | 1.09–1.30 | Z = 3.92, P < .001 | 87% (P < .001) |
| Diabetes | Cross-sectional | 3 | 9,667,143 | OR 1.21 | 1.11–1.32 | Z = 4.49, P < .001 | 84% (P = .002) |
| Diabetes | Cohort | 6 | 239,650 | HR 1.20 | 1.13–1.29 | Z = 5.38, P < .001 | 54% (P = .05) |
| Hyperlipidemia | Cohort | 3 | 42,524 | HR 1.21 | 1.13–1.29 | Z = 5.34, P < .001 | 51% (P = .13) |
| Obesity | Cross-sectional | 2 | 9,014,141 | OR 1.24 | 1.03–1.50 | Z = 2.27, P = .02 | 45% (P = .18) |
| Metabolic syndrome | Cross-sectional | 2 | 9,014,817 | OR 1.24 | 1.08–1.44 | Z = 2.95, P = .003 | 77% (P = .09) |

*OR, odds ratio; HR, hazard ratio; CI, confidence interval.*

## Discussion

### Principal findings

Twenty studies were included in the current review, screening 18,337 articles from four databases. Among the included studies, there were 11 cohort studies [3,15–24], 8 cross-sectional studies [1,2,25–30], and one mixed study [31] with both cohort and cross-sectional components. 18 studies [1–3,16–29,31] contributed to the meta-analysis. The Newcastle-Ottawa Scale (NOS) assessed the cohort studies and rated all as high quality. The modified Newcastle-Ottawa Scale assessed the cross-sectional studies and rated them as average to good quality. Visual assessment of the publication bias revealed minimal asymmetry in the funnel plot. Pooled evidence from cross-sectional and cohort studies showed that hysterectomy is associated with a 19% to 32% increased risk of hypertension. Pooled evidence from both cross-sectional studies and cohort studies reported that hysterectomy is associated with around a 20% increase in the odds of diabetes. Meta-analysis of cohort studies revealed that hysterectomy increased the risk of hyperlipidemia by 21%. Cross-sectional studies found that hysterectomy was associated with a 24% increase in the odds of both obesity and metabolic syndrome. In the largest study, Bi et al. reported that hysterectomy was associated with a 25% increased risk of MAFLD, analyzing over 9 million urban Chinese women [29].

### Interpretation of the findings

#### Premature ovarian failure as a unifying mechanism

Premature ovarian failure is the most widely proposed explanation for the observed associations between hysterectomy and cardiovascular risk factors [2,20,23,24,27,29]. The ovarian blood supply is derived from both the ovarian and uterine arteries via the utero-ovarian anastomosis. Ligation of the uterine vessels during hysterectomy may significantly reduce ovarian perfusion [31]. Therefore, even when the ovaries are preserved, hysterectomy may compromise ovarian function, which may explain why women who undergo hysterectomy with ovarian conservation experience earlier menopause compared to women with intact uteri [3,31]. Ovarian dysfunction leads to reduced levels of estrogen and other ovarian hormones with systemic metabolic consequences [17].

Adverse cardio-metabolic effects seen in premature ovarian failure are primarily due to estrogen deficiency. Estrogen contributes to glucose homeostasis by enhancing insulin sensitivity, promoting insulin secretion from pancreatic beta cells, facilitating glucose uptake via GLUT4 translocation to the plasma membrane, and suppressing hepatic gluconeogenesis [16,23]. The abrupt decline in estrogen following hysterectomy in premenopausal women may precipitate insulin resistance and increase diabetes risk. Estrogen plays a role in lipid metabolism by increasing HDL-cholesterol, decreasing LDL-cholesterol, and reducing lipoprotein (a) levels [15,30]. Estrogen also promotes blood pressure regulation by increasing nitric oxide bioavailability, inhibiting vascular smooth muscle proliferation, reducing angiotensin II receptor expression, and modulating the renin-angiotensin-aldosterone system [2,28]. The loss of these protective effects following hysterectomy may predispose women to hypertension. Estrogen also plays a role in obesity and metabolic syndrome through its effects on central appetite regulation, adipose tissue distribution, and adipocyte metabolism [28].

None of the included studies directly assessed postoperative ovarian function following hysterectomy with ovarian conservation. The physical presence of ovaries does not necessarily equate to functional ovaries, as the disruption of the utero-ovarian vascular supply may render conserved ovaries functionally compromised. This may explain why hysterectomy alone (without oophorectomy) consistently demonstrated positive associations with adverse metabolic outcomes. In addition, the relatively smaller sample sizes in the hysterectomy with bilateral oophorectomy groups limited the statistical power to detect the additional risk attributable to oophorectomy. Therefore, our meta-analytic approach combined the two groups to maximize statistical power while acknowledging that hysterectomy, regardless of ovarian conservation, appears to confer cardiovascular risk.

#### The cancellation effects of estrogen and androgen in bilateral oophorectomy

Some studies have shown that bilateral oophorectomy alone (without hysterectomy) does not increase cardiovascular risk factors and, in some cases, may even be protective [3,23,31]. These findings may reflect the dual hormonal effects of oophorectomy. Bilateral oophorectomy reduces not only estrogen production but also androgen production (particularly testosterone). The ovaries contribute approximately 50% of circulating testosterone in premenopausal women [3,23]. While estrogen deficiency increases the risk of diabetes and metabolic dysfunction, reduced androgen levels may mitigate or even reverse this risk [3,23]. In cases of hysterectomy alone with ovarian preservation, the extent of reduction in androgen and estrogen production due to vascular compromise may vary. Compromise of ovarian function with retention of androgen production can result in more pronounced metabolic effects than bilateral oophorectomy. Therefore, consider both postoperative estrogen and androgen levels when interpreting these findings.

#### Age-dependent effects and clinical significance

Multiple studies have shown that the association between hysterectomy and cardiovascular risk factors is more pronounced among younger women [2,3,19,20,23,24]. Madika et al. reported that the risk of hypertension increased with earlier age at hysterectomy, with a hazard ratio of 1.28 (95% CI: 1.13–1.45) for women under 40 years compared to 1.12 (95% CI: 1.03–1.22) for women over 50 years (p < 0.0001) [19]. Chiang et al. reported that women aged 30–39 years who underwent hysterectomy had a hazard ratio of 1.75 (95% CI: 1.27–2.41) for diabetes, compared with 1.33 (95% CI: 1.19–1.49) for women aged 40–49 years [24]. Increasing age itself is a well-recognized cardiovascular risk factor. The risk is relatively low in younger females [8]. When younger females undergo hysterectomy ± oophorectomy, they are compared with younger control groups, revealing the cardiovascular risk attributable to hysterectomy. However, increasing age may have overridden the cardiovascular risk attributable to hysterectomy in older females, showing no statistically significant difference between those who underwent surgery and control groups.

In younger women, the ovaries still produce significant amounts of estrogen and other hormones. The abrupt loss of these hormones may be more metabolically disruptive than in older women. Clinicians should be aware of the potential long-term cardiovascular consequences in women who undergo hysterectomy at younger ages. This is particularly important given that younger women may not typically be considered at high cardiovascular risk and may not receive routine screening for metabolic conditions.

#### The role of hormone replacement therapy and its documentation gaps

Hormone replacement therapy (HRT) is often prescribed to manage menopausal symptoms, particularly for younger women who experience surgical menopause. However, most of the included studies did not report data on HRT use, which may be a significant source of both confounding and effect modification [2,18,20,23,28,29,32]. Ouyang et al. addressed this issue most directly in their UK Biobank analysis. They found that HRT use in women who had undergone hysterectomy or bilateral oophorectomy was not associated with increased diabetes risk (hysterectomy alone: aHR1.01, 95% CI:0.84–1.21; hysterectomy with bilateral oophorectomy: aHR1.03, 95% CI:0.83–1.28) [23]. The lack of association may reflect exogenous hormones attenuating post-surgery hormonal changes. Several other studies adjusted for HRT use but did not provide detailed analyses of how HRT modified the associations [3,16,19,20,22].

#### The iron storage hypothesis

Premenopausal women experience regular menstrual blood loss, which leads to lower levels of stored iron (ferritin) and hemoglobin compared with men and postmenopausal women [20,25]. This relative iron deficiency has been proposed as a protective factor against cardiovascular disease. Excess iron promotes oxidative stress, catalyzing the production of highly reactive hydroxyl radicals that can oxidize LDL-cholesterol and promote atherosclerosis [20,25]. Hysterectomy permanently eliminates this physiological mechanism of iron loss, leading to an early increase in hematocrit and iron stores [20]. Elevated hematocrit increases whole-blood viscosity, leading to endothelial dysfunction and subsequent hypertension [20].

Yuk et al. found that the risk of hypertension following hysterectomy was significantly higher only in women whose hemoglobin increased by 2 g/dL or more after surgery (aHR1.34, 95% CI:1.22–1.42). The proportion of women with a hemoglobin increase of ≥2 g/dL was significantly higher in the hysterectomy group (35.7%) compared to the non-hysterectomy group (8.2%, p<0.001) [20]. These findings suggest that even when ovarian function is preserved, the cessation of menstrual bleeding may independently increase cardiovascular risk through iron-mediated pathways. However, as heavy menstrual bleeding is a common indication for hysterectomy, post-hysterectomy hemoglobin rise is an expected outcome. These findings may reflect confounding. Therefore, it is difficult to arrive at clinical decisions. To identify clinically meaningful differences in cardiovascular risk, RCTs should compare hysterectomy versus a levonorgestrel-releasing intrauterine system (LNG/IUS) in patients with heavy menstrual bleeding.

#### The uterus as an endocrine organ with functions beyond reproduction

The uterus may have endocrine or paracrine functions beyond its reproductive role, which can potentially mitigate cardiovascular risk factors. Hysterectomy may eliminate these protective functions even when the ovaries are conserved. Only a few studies have examined the association between hysterectomy performed after menopause and cardiovascular risk factors. The existing evidence is inadequate because of relatively small sample sizes [17,19,21,25,30,31]. Laughlin-Tommaso et al. found that women who underwent hysterectomy after age 50 did not have any added risk. However, their sample size in this age stratum was relatively small (271 vs 4,188), limiting the power to detect moderate associations [18]. The possibility that the uterus produces a substance (such as prostacyclin, prostaglandin, relaxin, various growth factors, or cytokines) that protects against cardiovascular disease even after menopause warrants further investigation [25].

#### The role of shared risk factors and confounding by indication

Some studies raise the concern that the same conditions that lead to hysterectomy may also increase cardiovascular risk factors, referred to as “confounding by indication”. The most common indications for hysterectomy, like uterine fibroids, endometriosis, and abnormal uterine bleeding, may themselves be associated with metabolic risk factors. Uterine fibroids are more common in women with obesity, hypertension, and diabetes [29]. The mechanisms linking fibroids to these conditions may involve shared inflammatory pathways, insulin resistance, and hormonal influences. Endometriosis is associated with chronic systemic inflammation, which may also increase the risk of hypertension through endothelial dysfunction and vascular inflammation [19]. Thus, women who undergo hysterectomy may already have a higher baseline cardiovascular risk profile that precedes the surgery.

Madika et al. found that endometriosis (aHR 1.19, 95% CI: 1.11–1.22) and fibroids (aHR 1.18, 95% CI: 1.13–1.22) were both independently associated with hypertension, even after adjustment for hysterectomy status and other confounders. Women with both hysterectomy and endometriosis had the highest risk (aHR 1.24, 95% CI: 1.11–1.38) [19]. Yan et al. found that genetically predicted hysterectomy was causally associated with hypertension (OR 1.205, 95% CI: 1.043–1.392) [2]. In the cross-sectional studies [1,26,28–31], the age at hysterectomy was not clearly documented for most participants, making it difficult to determine whether the surgery preceded the development of metabolic abnormalities or vice versa. This is a critical limitation because reverse causation, where pre-existing metabolic risk factors increase the likelihood of hysterectomy, cannot be ruled out.

#### Surveillance bias and health-seeking behavior

There is a possibility that women who undergo hysterectomy have more frequent contact with healthcare providers and are therefore more likely to be diagnosed with cardiovascular risk factors compared to women with intact uteri. This bias could inflate the apparent association between hysterectomy and metabolic outcomes by increasing the detection rate. Ding et al. found that the number of clinic visits and hospitalizations was higher in the hysterectomy group than in the comparison group, raising the possibility of surveillance bias [17]. Laughlin-Tommaso et al. showed that the median density of medical contacts during follow-up was 7.3 per year for women with hysterectomy and 6.2 per year for referent women, a modest but potentially meaningful difference [18]. Surveillance bias cannot be entirely ruled out, particularly in studies that rely on administrative databases, where diagnoses are recorded from clinical encounters rather than systematic screening.

### Strengths and limitations

To the best of our knowledge, this is the first comprehensive systematic review and meta-analysis specifically designed to evaluate the role of hysterectomy in modifying the main modifiable atherosclerotic cardiovascular disease (ASCVD) risk factors as defined by the European Society of Cardiology guidelines. The inclusion of studies from diverse populations, including American, European, Korean, Taiwanese, Chinese, Indian, and multi-ethnic cohorts, enhances the generalizability of the findings. The inclusion of high-quality studies strengthened the evidence.

Despite these strengths, several limitations remain. Heterogeneity across studies was substantial, so the pooled estimates should be interpreted with caution. We were unable to perform subgroup analyses comparing hysterectomy alone with hysterectomy with bilateral oophorectomy, or to assess age at hysterectomy, because few studies provided separate estimates. The reliance on self-reported hysterectomy status in several studies may introduce bias [2,16]. Outcome definitions and confounder adjustment varied across studies, affecting comparability. The lack of data on hormone replacement therapy use is a significant limitation. The temporality of the association cannot be established with cross-sectional studies, making it difficult to rule out reverse causation. The studies did not explicitly report the surgical approach to hysterectomy (abdominal, vaginal, or laparoscopic), which may have modified the associations discussed.

### Implications

The mechanisms linking hysterectomy to cardiovascular risk are not fully understood and warrant further research. Studies directly measuring ovarian function, hormone levels, and iron status following hysterectomy would help elucidate the relative contributions of these pathways. Precise documentation of HRT use after hysterectomy is essential to understand its role in modifying cardiovascular risk. Prospective cohort studies with long follow-up data are needed to establish temporal relationships and quantify the absolute risk associated with hysterectomy at different ages. Randomized controlled trials comparing hysterectomy with alternatives (such as uterine artery embolization) would provide the strongest evidence of its causal effects.

Mendelian randomization studies with larger sample sizes and more comprehensive genetic data could provide additional evidence of causality. While hysterectomy provides important benefits for women with symptomatic gynecological conditions, the long-term metabolic consequences should be considered in shared decision-making. For women who undergo hysterectomy, particularly at a young age, cardiovascular risk factor screening should be initiated early and continued throughout the lifespan.

## Conclusions

This systematic review and meta-analysis provide comprehensive evidence that hysterectomy is associated with increased risk of hypertension, diabetes, hyperlipidemia, obesity, and metabolic syndrome, which are key modifiable atherosclerotic cardiovascular disease risk factors. Multiple pathophysiological mechanisms, including premature ovarian failure, the iron storage hypothesis, cancellation effects of ovarian hormones, and the paracrine or endocrine role of the uterus beyond reproduction, are proposed to describe the biological plausibility of these associations. There is a need for active cardiovascular risk assessment in women who undergo hysterectomy, particularly at a young age. Therefore, hysterectomy should not be viewed merely as a gynecological procedure, but rather as a significant modifier of cardiovascular risk. Future research should further elucidate the mechanisms underlying these associations.

## Data Availability

All relevant data are within the manuscript and its Supporting Information files.

## Acknowledgments

During the preparation of this work, the authors used Grammarly Premium and Claude for language improvement only. The authors ensured the accuracy and validity of all manuscript data. No AI tools were used to generate data, results, or concepts. The authors reviewed and edited the content as needed and take full responsibility for the content of the published article.

## Supporting information

**S1 File. Search strategy**. Database-specific, line-by-line search strategies for PubMed, Cochrane Library, Embase, and ScienceDirect, searched from inception to March 10, 2026.

**S2 File. PRISMA 2020 checklist**. Completed PRISMA 2020 checklist for this systematic review and meta-analysis.

## Notes

### Competing Interest Statement

The authors have declared no competing interest.

