## Supplementary File 1 for "Decision for hysterectomy as a modifiable cardiovascular risk factor: A systematic review and meta-analysis"

**Search strategy**

We extracted Title-Abstract files in the formats of csv, ris, bib, and .txt from the following databases to be exported and included in Rayyan. All databases were searched from inception to 10/03/2026.

**Cochrane Library**

We conducted an advanced search on 10/03/2026. We typed each search term under ‘search manager’ in the Cochrane database. We have reported results for each search term separately. Then, we combined the search terms with the Boolean expression OR, resulting in 757 articles.

ID           Search                Hits

#1          Hysterectomy AND cardiovascular disease **150**

#2          Hysterectomy AND diabetes **276**

#3          Hysterectomy AND coronary artery disease **36**

#4          Hysterectomy AND ischemic heart disease **31**

#5          Hysterectomy AND hypertension **169**

#6          Hysterectomy AND dyslipidemia **19**

#7          Hysterectomy AND hyperlipidemia **13**

#8          Hysterectomy AND metabolic syndrome **18**

#9          Hysterectomy AND metabolic morbidity **03**

#10       Hysterectomy AND heart disease **194**

#11       Hysterectomy AND cardiac morbidity **22**

#12  #1 OR #2 OR #3 OR #4 OR #5 OR #6 OR #7 OR #8 OR #9 OR #10 OR #11 **757**

**Embase**

We conducted an advanced search on 10/03/2026. The results showed 7387 articles. The detailed steps of the search strategy are shown below.

ID           Search                Hits

#1          Hysterectomy AND cardiovascular disease **927**

#2          Hysterectomy AND diabetes **3590**

#3          Hysterectomy AND coronary artery disease **298**

#4          Hysterectomy AND ischemic heart disease **367**

#5          Hysterectomy AND hypertension **4251**

#6          Hysterectomy AND dyslipidemia **248**

#7          Hysterectomy AND hyperlipidemia **342**

#8          Hysterectomy AND metabolic syndrome **140**

#9          Hysterectomy AND metabolic morbidity **01**

#10       Hysterectomy AND heart disease **887**

#11       Hysterectomy AND cardiac morbidity **07**

#12  #1 OR #2 OR #3 OR #4 OR #5 OR #6 OR #7 OR #8 OR #9 OR #10 OR #11 **7387**

**PubMed**

We conducted an advanced search on 10/03/2026. The results showed 2230 articles. We have introduced no filters. The search strings are mentioned below.

ID           Search                Hits

#1          Hysterectomy AND cardiovascular disease **1240**

#2          Hysterectomy AND diabetes **955**

#3          Hysterectomy AND coronary artery disease **97**

#4          Hysterectomy AND ischemic heart disease **244**

#5          Hysterectomy AND hypertension **854**

#6          Hysterectomy AND dyslipidemia **54**

#7          Hysterectomy AND hyperlipidemia **52**

#8          Hysterectomy AND metabolic syndrome **57**

#9          Hysterectomy AND metabolic morbidity **655**

#10       Hysterectomy AND heart disease **911**

#11       Hysterectomy AND cardiac morbidity **479**

#12  #1 OR #2 OR #3 OR #4 OR #5 OR #6 OR #7 OR #8 OR #9 OR #10 OR #11 **2230**

**ScienceDirect**

We conducted an advanced search on 10/03/2026 using the following keywords combined with Boolean expressions. However, Science Direct allows a maximum of eight Boolean expressions only. Therefore, we modified the search terms and Boolean expressions used for other databases, excluding several terms. Under advanced search, we typed the following search terms combined with Boolean expressions in the section: ‘Find articles with these terms.’ We used two filters: Article type (Research articles) and language (English). The results showed 7963 articles.

(Hysterectomy) AND ((Cardiovascular disease) OR (Diabetes) OR (Hypertension) OR (Dyslipidemia) OR (Hyperlipidemia) OR (Metabolic syndrome) OR (Metabolic morbidity)) **7963**
