## Supplementary File 2 for "Decision for hysterectomy as a modifiable cardiovascular risk factor: A systematic review and meta-analysis"

| **Section and Topic** | **Item #** | **Checklist item** | **Location where item is reported** |
| --- | --- | --- | --- |
| **TITLE** | | |  |
| Title | 1 | Identify the report as a systematic review. | Title |
| **ABSTRACT** | | |  |
| Abstract | 2 | See the PRISMA 2020 for Abstracts checklist. | Abstract |
| **INTRODUCTION** | | |  |
| Rationale | 3 | Describe the rationale for the review in the context of existing knowledge. | Introduction |
| Objectives | 4 | Provide an explicit statement of the objective(s) or question(s) the review addresses. | Introduction, Objective (see also Abstract) |
| **METHODS** | | |  |
| Eligibility criteria | 5 | Specify the inclusion and exclusion criteria for the review and how studies were grouped for the syntheses. | Materials and methods, Eligibility criteria |
| Information sources | 6 | Specify all databases, registers, websites, organisations, reference lists and other sources searched or consulted to identify studies. Specify the date when each source was last searched or consulted. | Materials and methods, Information sources (see also Abstract, Data sources) |
| Search strategy | 7 | Present the full search strategies for all databases, registers and websites, including any filters and limits used. | Materials and methods, Search strategy; full search strings provided in S1 File |
| Selection process | 8 | Specify the methods used to decide whether a study met the inclusion criteria of the review, including how many reviewers screened each record and each report retrieved, whether they worked independently, and if applicable, details of automation tools used in the process. | Materials and methods, Study selection |
| Data collection process | 9 | Specify the methods used to collect data from reports, including how many reviewers collected data from each report, whether they worked independently, any processes for obtaining or confirming data from study investigators, and if applicable, details of automation tools used in the process. | Materials and methods, Data extraction |
| Data items | 10a | List and define all outcomes for which data were sought. Specify whether all results that were compatible with each outcome domain in each study were sought (e.g. for all measures, time points, analyses), and if not, the methods used to decide which results to collect. | Materials and methods, Eligibility criteria; Introduction, Objective (outcomes defined per ESC modifiable ASCVD risk factors) |
|  | 10b | List and define all other variables for which data were sought (e.g. participant and intervention characteristics, funding sources). Describe any assumptions made about any missing or unclear information. | Materials and methods, Data extraction; study characteristics extracted are shown in Tables 1-4 |
| Study risk of bias assessment | 11 | Specify the methods used to assess risk of bias in the included studies, including details of the tool(s) used, how many reviewers assessed each study and whether they worked independently, and if applicable, details of automation tools used in the process. | Materials and methods, Risk of bias and quality assessment |
| Effect measures | 12 | Specify for each outcome the effect measure(s) (e.g. risk ratio, mean difference) used in the synthesis or presentation of results. | Materials and methods, Data synthesis |
| Synthesis methods | 13a | Describe the processes used to decide which studies were eligible for each synthesis (e.g. tabulating the study intervention characteristics and comparing against the planned groups for each synthesis (item #5)). | Materials and methods, Data synthesis |
|  | 13b | Describe any methods required to prepare the data for presentation or synthesis, such as handling of missing summary statistics, or data conversions. | Materials and methods, Data synthesis |
|  | 13c | Describe any methods used to tabulate or visually display results of individual studies and syntheses. | Materials and methods, Data synthesis; Table 7; Figs 2-8 |
|  | 13d | Describe any methods used to synthesize results and provide a rationale for the choice(s). If meta-analysis was performed, describe the model(s), method(s) to identify the presence and extent of statistical heterogeneity, and software package(s) used. | Materials and methods, Data synthesis |
|  | 13e | Describe any methods used to explore possible causes of heterogeneity among study results (e.g. subgroup analysis, meta-regression). | Materials and methods, Data synthesis (stratification by study design and effect measure as the pre-specified approach to heterogeneity) |
|  | 13f | Describe any sensitivity analyses conducted to assess robustness of the synthesized results. | Materials and methods, Data synthesis |
| Reporting bias assessment | 14 | Describe any methods used to assess risk of bias due to missing results in a synthesis (arising from reporting biases). | Materials and methods, Data synthesis (funnel plot asymmetry); Egger's test not performed, as noted in Results, Risk of bias and quality assessment |
| Certainty assessment | 15 | Describe any methods used to assess certainty (or confidence) in the body of evidence for an outcome. | Not reported — a formal certainty/GRADE assessment was not performed for this review |
| **RESULTS** | | |  |
| Study selection | 16a | Describe the results of the search and selection process, from the number of records identified in the search to the number of studies included in the review, ideally using a flow diagram. | Results, Study characteristics; Fig 1 (PRISMA 2020 flow diagram) |
|  | 16b | Cite studies that might appear to meet the inclusion criteria, but which were excluded, and explain why they were excluded. | Fig 1 reports reasons for full-text exclusion (ongoing study, n=2; incomplete data, n=6); individual excluded studies are not cited by name in the main text |
| Study characteristics | 17 | Cite each included study and present its characteristics. | Results, Study characteristics; Tables 1-4 |
| Risk of bias in studies | 18 | Present assessments of risk of bias for each included study. | Results, Risk of bias and quality assessment; Tables 5 and 6 |
| Results of individual studies | 19 | For all outcomes, present, for each study: (a) summary statistics for each group (where appropriate) and (b) an effect estimate and its precision (e.g. confidence/credible interval), ideally using structured tables or plots. | Tables 1-4 (study-level exposed/unexposed summary data and effect estimates); Figs 2-8 (forest plots) |
| Results of syntheses | 20a | For each synthesis, briefly summarise the characteristics and risk of bias among contributing studies. | Results (subsections by outcome: hypertension, diabetes, lipid profile, obesity, metabolic syndrome) |
|  | 20b | Present results of all statistical syntheses conducted. If meta-analysis was done, present for each the summary estimate and its precision (e.g. confidence/credible interval) and measures of statistical heterogeneity. If comparing groups, describe the direction of the effect. | Results; Table 7; Figs 2-8 |
|  | 20c | Present results of all investigations of possible causes of heterogeneity among study results. | Results, Synthesis of results, Sensitivity analysis |
|  | 20d | Present results of all sensitivity analyses conducted to assess the robustness of the synthesized results. | Results, Synthesis of results, Sensitivity analysis |
| Reporting biases | 21 | Present assessments of risk of bias due to missing results (arising from reporting biases) for each synthesis assessed. | Results, Risk of bias and quality assessment |
| Certainty of evidence | 22 | Present assessments of certainty (or confidence) in the body of evidence for each outcome assessed. | Not reported — a formal certainty/GRADE assessment was not performed for this review |
| **DISCUSSION** | | |  |
| Discussion | 23a | Provide a general interpretation of the results in the context of other evidence. | Discussion, Principal findings; Interpretation of the findings |
|  | 23b | Discuss any limitations of the evidence included in the review. | Discussion, Strengths and limitations |
|  | 23c | Discuss any limitations of the review processes used. | Discussion, Strengths and limitations (review-process-specific limitations, e.g. database coverage and English-language restriction, are not separately discussed) |
|  | 23d | Discuss implications of the results for practice, policy, and future research. | Discussion, Implications |
| **OTHER INFORMATION** | | |  |
| Registration and protocol | 24a | Provide registration information for the review, including register name and registration number, or state that the review was not registered. | Materials and methods, Study selection (PROSPERO CRD420251241667) |
|  | 24b | Indicate where the review protocol can be accessed, or state that a protocol was not prepared. | PROSPERO registration record: https://www.crd.york.ac.uk/PROSPERO/view/CRD420251241667 |
|  | 24c | Describe and explain any amendments to information provided at registration or in the protocol. | Not reported — no amendments to the registered protocol are described |
| Support | 25 | Describe sources of financial or non-financial support for the review, and the role of the funders or sponsors in the review. | Funding Statement (entered in the PLOS ONE submission form): no specific funding received |
| Competing interests | 26 | Declare any competing interests of review authors. | Competing Interests statement (entered in the PLOS ONE submission form): none declared |
| Availability of data, code and other materials | 27 | Report which of the following are publicly available and where they can be found: template data collection forms; data extracted from included studies; data used for all analyses; analytic code; any other materials used in the review. | Data Availability Statement (entered in the PLOS ONE submission form); extracted study data are in Tables 1-7 and Figs 2-8, and the search strategy is in S1 File |

*From:*  Page MJ, McKenzie JE, Bossuyt PM, Boutron I, Hoffmann TC, Mulrow CD, et al. The PRISMA 2020 statement: an updated guideline for reporting systematic reviews. BMJ 2021;372:n71. doi: 10.1136/bmj.n71. This work is licensed under CC BY 4.0. To view a copy of this license, visit <https://creativecommons.org/licenses/by/4.0/>
